# Rapid Decline of Mpox Antibody Responses Following MVA-BN Vaccination

**DOI:** 10.1101/2024.09.10.24313399

**Authors:** Ai-ris Y. Collier, Katherine McMahan, Catherine Jacob-Dolan, Jinyan Liu, Erica N Borducchi, Bernard Moss, Dan H. Barouch

**Affiliations:** Beth Israel Deaconess Medical Center, Harvard Medical School, Boston, MA, USA; Laboratory of Viral Diseases, National Institute of Allergy and Infectious Diseases, National Institutes of Health, Bethesda, MD, USA

## Abstract

The replication-incompetent modified vaccinia Ankara-Bavarian Nordic vaccine (MVA-BN; Jynneos) was deployed during the 2022 clade IIb mpox outbreak. On August 14, 2024, the World Health Organization declared the mpox clade Ib outbreak in the Democratic Republic of the Congo a public health emergency of international concern, which has raised the question about the durability of vaccine immunity after MVA-BN vaccination. In this study, we show that the MVA-BN vaccine generated mpox serum antibody responses that largely waned after 6-12 months.

## Introduction

The orthopoxvirus genus consists of 12 viruses including smallpox (variola), mpox, and vaccinia viruses. The replication-incompetent modified vaccinia Ankara-Bavarian Nordic vaccine (MVA-BN; Jynneos) was developed as part of the U.S. Strategic National Stockpile program for deployment in event of smallpox outbreak, and the Centers for Disease Control and Prevention recommended MVA-BN vaccination for mpox infection during the 2022 clade IIb mpox outbreak. Due to limited supply, the recommended MVA-BN administration was modified from 0.5 mL by the subcutaneous (SC) route to 0.1 mL by the intradermal (ID) route. On August 14, 2024, the World Health Organization declared the mpox clade Ib outbreak in the Democratic Republic of the Congo a public health emergency of international concern,^1^ which has raised the question about the durability of vaccine immunity and reinfection risk^2^ after MVA-BN vaccination.

## Methods

We performed an observational study in 45 adults who received the MVA-BN (Jynneos) vaccine or had confirmed diagnosis of mpox infection at Beth Israel Deaconess Medical Center (BIDMC) in Boston, MA (**Table 1**). The BIDMC institutional review board approved this study. All participants provided informed consent. We assessed serum antibody and T cell responses for 12 months following either 2-dose or 1-dose MVA-BN vaccination delivered by either the SC or ID route.

**Table 1.** Participant characteristics by MVA-BN vaccination or mpox infection groups.

|  | **All**<br>**N=45** | **1 dose (ID or SC)**<br>**N=26\*** | **2 dose (2 SC, 2 ID,**<br>**or SC+ID)**<br>**N=22\*** | **Convalescent**<br>**N=3** |
| --- | --- | --- | --- | --- |
| Age (years) at vaccine, median [range] | 25 [20-66] | 25 [22-53] | 25 [20-66] | 30 [26-35] |
| Sex at birth (female) | 24 (53) | 17 (65) | 11 (50) | 0 |
| Race |  |  |  |  |
| White | 27 (60) | 16 (62) | 13 (59) | 2 (66) |
| Black | 5 (11) | 3 (12) | 1 (5) | 1 (33) |
| Asian | 9 (20) | 6 (23) | 5 (23) | 0 |
| Native Hawaiian/Pacific Islander | 1 (2) | 0 | 1 (5) | 0 |
| Other | 3 (7) | 1 (4) | 2 (9) | 0 |
| Ethnicity |  |  |  |  |
| Hispanic or Latino | 5 (11) | 3 (12) | 1 (5) | 1 (33) |
| Not Hispanic or Latino | 40 (89) | 23 (88) | 21 (95) | 2 (66) |
| Medical history |  |  |  |  |
| Obesity | 3 (7) | 1 (4) | 3 (14) | 0 |
| Hypertension | 3 (7) | 2 (8) | 1 (5) | 0 |
| Asthma | 5 (11) | 4 (15) | 2 (9) | 0 |
| Living with HIV | 4 (9) | 2 (8) | 2 (9) | 1 (33) |
| MVA-BN administered SC | 11 (24) | 4 (15) | 7 (32) | N/A |
| MVA-BN administered ID | 27 (60) | 22 (85) | 8 (36) | N/A |
| MVA-BN administered SC+ID | 7 (16) | N/A | 7 (32) | N/A |
| Days from last vaccine/infection (peak) | 21 (17, 28) | 22 (19, 30) | 20 (16, 28) | 38 (38, 38) |
| Days from last vaccine/infection (3 months) | 94 (90, 98) | 92 (89, 96) | 96 (91, 100) | 95 (91, 99) |
| Days from last vaccine/infection (6 months) | 184 (181, 189) | 183 (180, 189) | 182 (182, 196) | 171 (143, 186) |
| Days from last vaccine/infection (9 months) | 277 (274, 281) | 278 (274, 282) | 276 (272, 281) | 287 (275, 311) |
| Days from last vaccine/infection (12 months) | 370 (365, 377) | 370 (366, 378) | 367 (365, 371) | 387 (387, 438) |
| Days between vaccine doses | N/A | N/A | 30 (28, 54) | N/A |

ID= intradermal administration, SC= subcutaneous administration, MVA-BN= modified vaccinia Ankara-Bavarian Nordic (Jynneos) vaccine, N/A= not applicable
Data presented as number (percent) or median (interquartile range), unless otherwise specified.
None had immunosuppression, no diabetes history.
\*Six individuals contributed to early 1-dose and later 2-dose timepoints due to long interval prior to receiving second dose.
\*\*Reporting days following infection rather than vaccination

## Results

Median binding antibody ELISA titers to mpox M1R, B6R, A35R, A29L, H3L antigens were 28, 25, 25, 27, 27 at baseline, respectively, and peaked at 112, 384, 85, 29, 76 at week 3 following 2 doses of MVA-BN but then declined to 38, 82, 32, 25, 31 at 12 months (**Fig. 1A**). In contrast, in participants who received only 1 dose of MVA-BN, the median binding antibody ELISA titers to mpox M1R, B6R, A35R, A29L, H3L antigens peaked at 45, 90, 32, 31, 28 at week 3 but then declined to 33, 43, 30, 25, 28 at 12 months (**Fig. 1B**). Mpox serum neutralizing antibody (NAb) titers were detectable in only a few participants following 2-dose or 1-dose MVA-BN vaccines (median titers 11 and 9.5, respectively) at 3 months. High titers of mpox NAbs (median titer 965) were detected at 3 months following natural infection and persisted at 9 months post-infection (median titer 284; **Fig. 1C**).

**Figure 1.**
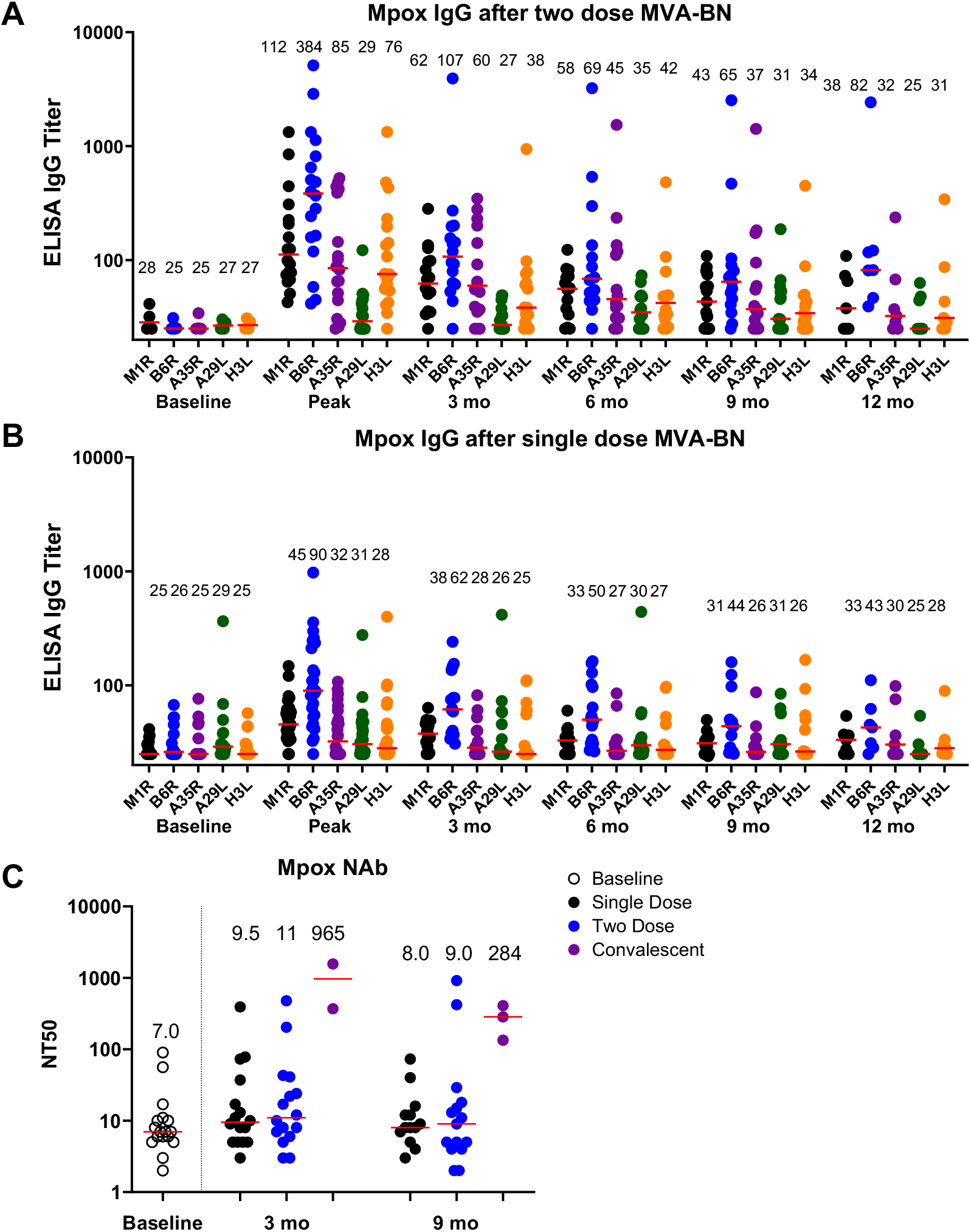
Serum mpox binding and neutralizing antibody responses following 2-dose and 1-dose MVA-BN vaccination

Low peripheral IFN-γ CD4+ and CD8+ T cell responses were detected to vaccinia-infected target cells by intracellular cytokine staining assays at 9 months following 2-dose and 1-dose MVA-BN vaccination (median CD4 responses 0.024%, 0.016%, respectively; median CD8 responses 0.053%, 0.013%, respectively) but were not detected by mpox peptide-specific ELISPOT assays (data not shown).

## Discussion

MVA-BN provided 66% efficacy as a 2-dose regimen and 36% efficacy as a 1-dose regimen at peak immunity during the 2022 mpox outbreak.^3^ Our data demonstrates that MVA-BN vaccination generated mpox antibodies that largely waned after 6-12 months.^4^ Specifically, in participants who received the 2-dose MVA-BN vaccine, mpox antibody responses at 12 months were comparable or lower than peak antibody responses in people who received the 1-dose MVA-BN vaccine that provided limited protection. Serum antibody titers following vaccination have been shown to correlate with protection against mpox challenge in nonhuman primates^56^, whereas CD4+ and CD8+ T cell responses did not correlate with protection, suggesting the potential relevance of serum antibody titers following MVA-BN vaccination in humans. Moreover, a cluster of mpox infections was reported in 2023 in vaccinated humans with waning immunity.^2^ Taken together, these data suggest that protective immunity may be waning in individuals who were vaccinated with MVA-BN in 2022 and that boosting may be required to maintain robust levels of protective immunity. Strategies are needed to improve the durability of mpox vaccines.

## Data Availability

All data produced in the present work are contained in the manuscript

## Correspondence

Correspondence and requests for materials should be addressed to D.H.B.

## Author Contributions

AYC: concept and design, acquisition of data, data analysis, interpretation of data, drafting of the manuscript, critical revision of the manuscript, statistical analysis, administrative support, technical support, material support, supervision

KAM: acquisition of data, interpretation of data, critical revision of the manuscript, technical support

CJD: data analysis, interpretation of data, administrative support, technical support, critical revision of the manuscript

JL: acquisition of data, data analysis, critical revision of the manuscript, technical support ENB: acquisition of data, critical revision of the manuscript, administrative support, technical support

BM: acquisition of data, data analysis, interpretation of data, critical revision of the manuscript, material support

DHB: concept and design, data analysis, interpretation of data, drafting of the manuscript, critical revision of the manuscript, obtained funding, administrative support, technical support, material support, supervision

## Acknowledgements

We thank Lydia Gallup, M.S.N., R.N., Siline Thai, B.S., Eleanor Schonberg, B.S., Marjorie Rowe, B.S., Tochi Anioke, B.S., Amelia Hoyt, B.S., Orion Barnett, B.S., Juliana Pereira, B.S., Audrey Mutoni, B.S., Bridget Wixted, B.S., Krishna Shah, B.S., Bismark Acquah, M.S., Kristin Gotthardt, B.S., Reed Boduch, B.S., and Michelle A. Lifton, M.S., Maxinne Ignacio, B.S., and Catherine Cotter, M.S. for participant recruitment, sample collection, and technical assistance.

## Supplementary Methods

### Study Population

A specimen biorepository at Beth Israel Deaconess Medical Center (BIDMC) obtained peripheral blood and clinical data from adults who received the modified vaccinia Ankara-Bavarian Nordic (MVA-BN; Jynneos) vaccine or had confirmed diagnosis of mpox infection. The BIDMC institutional review board approved this study (2022P000577). All participants provided informed consent.

### Enzyme-linked immunosorbent assay (ELISA)

Mpox A29L, A35R, B6R, H3L, and M1R binding antibodies in serum were assessed by ELISA in BSL2+ containment as previously described.^1,2^ Ninety-six–well plates were coated with mpox A29L, A35R, B6R, H3L, or M1R protein (1 μg/ml; Sino Biological) in 1× Dulbecco phosphate-buffered saline (DPBS) and incubated at 4°C overnight. After incubation, plates were washed once with wash buffer (0.05% Tween 20 in 1× DPBS) and blocked with 350 μl of casein block solution per well for 2 to 3 hours at room temperature. After incubation, block solution was discarded, and plates were blotted dry. Serial dilutions of heat-inactivated serum diluted in casein block were added to wells, and plates were incubated for 1 hour at room temperature, before three more washes, and a 1-hour incubation with a 1:4000 dilution of anti–human immunoglobulin G (IgG) horseradish peroxidase (Invitrogen, ThermoFisher Scientific) at room temperature in the dark. Plates were washed three times, and 100 μl of SeraCare KPL TMB SureBlue Start solution was added to each well; plate development was halted by adding 100 μl of SeraCare KPL TMB Stop solution per well. The absorbance at 450 nm, with a reference at 650 nm, was recorded with a VersaMax microplate reader (Molecular Devices). For each sample, the ELISA endpoint titer was calculated using a four parameter logistic curve fit to calculate the reciprocal serum dilution that yields a corrected absorbance value (450 to 650 nm) of 0.2. Interpolated end point titers are reported.

### Mpox Virus Neutralizing Antibody Assay

A rapid, sensitive, and quantitative 96-well plate semi-automated, flow cytometric assay was carried out using Mpox Z-1979 expressing Aequorea coerulescens green fluorescent protein (AcGFP) as previously described.^3^ Several twofold dilutions of heat-inactivated immune serum (56°C for 30 min) from individual nonhuman primates were prepared in 96-well, round-bottom polypropylene plates using spinner modified minimum essential medium containing 2% FBS (Spinner-2%) and 2.5 % guinea pig complement (fresh frozen, Rockland). In BSL3 containment, approximately 2.5 × 104 PFU of Mpox-Z79 green fluorescent protein (GFP)–expressing virus was added to each well, and plates were incubated at 37°C for 1 hour. After incubation, 105 HeLa S3 (RRID: CVCL_0058) cells were pipetted into each well, and plates were incubated for an additional 16 to 18 hours at 37°C. The cells were fixed in 2% paraformaldehyde to inactivate virus, and GFP expression was measured and quantitated at BSL1 using a fluorescence-activated cell sorting (FACS) Canto II flow cytometer and FlowJo software (BD Biosciences). Half-maximal neutralization titer values were calculated using Prism software (GraphPad) to plot dose-response curves, normalized using the average of no-virus wells as 100% neutralization and no-serum wells as 0%.

### Vaccinia virus-specific intracellular cytokine staining (ICS) assay

10^6^ PBMCs/well were re-suspended in 100 *µ*L of R10 media supplemented with CD28 (1 *µ*g/mL) and CD49d (1 *µ*g/mL) monoclonal antibodies. Each sample was assessed with mock (100*µ*L of R10 plus 0.5% DMSO; background control), 1 × 10^7^ virial particle of the Western Reserve strain of vaccinia virus, or 10 pg/mL phorbol myristate acetate (PMA) and 1 *µ*g/mL ionomycin (Sigma-Aldrich) (100*µ*L; positive control) and incubated at 37°C for 3 hours. After incubation, 0.25 *µ*L of GolgiStop and 0.25 *µ*L of GolgiPlug in 50 *µ*L of R10 was added to each well and incubated at 37°C for 6 h and then held at 4°C overnight. The next day, the cells were washed twice with DPBS, stained with Aqua live/dead dye for 10 mins and then stained with predetermined titers of mAbs against CD279 (clone EH12.1, BB700), CD4 (clone L200, BV711), CD27 (clone M-T271, BUV563), CD8 (clone SK1, BUV805), CD45RA (clone 5H9, APC H7) for 30 min. Cells were then washed twice with 2% FBS/DPBS buffer and incubated for 15 min with 200 *µ*L of BD CytoFix/CytoPerm Fixation/Permeabilization solution. Cells were washed twice with 1X Perm Wash buffer (BD Perm/WashTM Buffer 10X in the CytoFix/CytoPerm Fixation/Permeabilization kit diluted with MilliQ water and passed through 0.22*µ*m filter) and stained intracellularly with monoclonal antibodies against Ki67 (clone B56, BB515), IL21 (clone 3A3-N2.1, PE), CD69 (clone TP1.55.3, ECD), IL10 (clone JES3-9D7, PE CY7), IL13 (clone JES10-5A2, BV421), IL4 (clone MP4-25D2, BV605), TNF-α (clone Mab11, BV650), IL17 (clone N49-653, BV750), IFN-γ (clone B27; BUV395), IL2 (clone MQ1-17H12, BUV737), IL6 (clone MQ2-13A5, APC), and CD3 (clone SP34.2, Alexa 700) for 30 min. Cells were washed twice with 1X Perm Wash buffer and fixed with 250*µ*L of freshly prepared 1.5% formaldehyde. Fixed cells were transferred to 96-well round bottom plate and analyzed by BD FACSymphonyTM system.

## References

1. WHO Director-General declares mpox outbreak a public health emergency of international concern. Accessed August 21, 2024. https://www.who.int/news/item/14-08-2024-who-director-general-declares-mpox-outbreak-a-public-health-emergency-of-international-concern

2. Faherty EAG. Notes from the Field: Emergence of an Mpox Cluster Primarily Affecting Persons Previously Vaccinated Against Mpox — Chicago, Illinois, March 18–June 12, 2023. MMWR Morb Mortal Wkly Rep. 2023;72. doi:10.15585/mmwr.mm7225a6

3. Deputy NP, Deckert J, Chard AN, et al. Vaccine Effectiveness of JYNNEOS against Mpox Disease in the United States. N Engl J Med. Published online May 18, 2023. doi:10.1056/NEJMoa2215201

4. Berry MT, Khan SR, Schlub TE, et al. Predicting vaccine effectiveness for mpox. Nat Commun. 2024;15(1):3856. doi:10.1038/s41467-024-48180-w

5. Jacob-Dolan C, Ty D, Hope D, et al. Comparison of the immunogenicity and protective efficacy of ACAM2000, MVA, and vectored subunit vaccines for Mpox in rhesus macaques. Sci Transl Med. 2024;16(740):eadl4317. doi:10.1126/scitranslmed.adl4317

6. Mucker EM, Freyn AW, Bixler SL, et al. Comparison of protection against mpox following mRNA or modified vaccinia Ankara vaccination in nonhuman primates. Cell. Published online September 4, 2024. doi:10.1016/j.cell.2024.08.043

## Supplementary References

1. Aid M, Sciacca M, McMahan K, et al. Mpox infection protects against re-challenge in rhesus macaques. Cell. 2023;186(21):4652-4661.e13. doi:10.1016/j.cell.2023.08.023

2. Jacob-Dolan C, Ty D, Hope D, et al. Comparison of the immunogenicity and protective efficacy of ACAM2000, MVA, and vectored subunit vaccines for Mpox in rhesus macaques. Sci Transl Med. 2024;16(740):eadl4317. doi:10.1126/scitranslmed.adl4317

3. Freyn AW, Atyeo C, Earl PL, et al. An mpox virus mRNA-lipid nanoparticle vaccine confers protection against lethal orthopoxviral challenge. Sci Transl Med. 2023;15(716):eadg3540. doi:10.1126/scitranslmed.adg3540

